# Safety and Effectiveness of SA58 Nasal Spray against COVID-19 Infection in Medical Personnel:An Open-label, Blank-controlled Study

**DOI:** 10.1101/2022.12.27.22283698

**Authors:** Shujie Si, Canrui Jin, Jianping Li, Yunlong Cao, Biao Kan, Feng Xue, Xiaoliang Sunney Xie, Liang Fang, Gang Zeng, Shuo Zhang, Yaling Hu, Xiaoping Dong

## Abstract

Approved COVID-19 vaccines to date have limited effectiveness in protecting infection and blocking transmission. A nasal spray of broad-spectrum antibody against COVID-19 (SA58 Nasal Spray) has recently been developed by Sinovac Life Sciences Co., Ltd.. From October 31 to November 30, 2022, an open-label, blank controlled study on the SA58 Nasal Spray against COVID-19 infection was conducted with the medical personnel working in the designated COVID-19 hospitals and Fangcang shelter hospitals (alternate care sites) of COVID-19 cases in Hohhot city, the Inner Mongolia Autonomous Region. A total of 6662 medical personnel were involved in this study: 3368 used SA58 Nasal Spray from the drug group, and 3294 not used from blank control group. The medication was self-administered intranasally 1∼2 times per day with an interval of 6 hours for 30 days.. The safety results indicated that the SA58 Nasal Spray was well tolerant. The incidence of adverse events (AEs) was 28.6% (497/1736), and the majority of the AEs were mild and from administrative site. 135 COVID-19 cases were identified for SARS-CoV-2 by RT-PCR during the 30-day observation. The cumulative incidence of COVID-19 in the drug group and the control group were 0.026% and 0.116%, respectively. The effectiveness of the SA58 Nasal Spray for preventing COVID-19 infection among medical personnel was evaluated as 77.7% (95% CI: 52.2% - 89.6%). In conclusion, the SA58 Nasal Spray is well-tolerant and highly effective against COVID-19 infection.

## Introduction

The COVID-19 pandemic has had a severe impact on public health, the economy and society worldwide, and has led to 646,266,987 COVID-19 cases and 6,636,278 deaths globally by the middle of December 2022^[1]^. One of the major biological features of SARS-CoV-2 is the continuous emergence of the mutants or viral variants that alter communicability, clinical outcomes, and immune evasion. According to the WHO classification, several variants of concern (VOCs) for SARS-CoV-2 were identified, including Alpha, Beta, Gamma, Delta and Omicron VOCs. Since the emergence of Omicron VOC in South Africa in the end of 2021, it rapidly spread across the globe and replaced previous viral strains as the predominant strain. Furthermore, Omicron VOC is still evolving and mutating, which has had more than 500 sub-variants or sublineages based on the PANGO lineage^[2, 3]^.

Billions of people have been vaccinated with different types of COVID-19 vaccine worldwide. Although they have shown great efficacy in protecting against severe/critical cases and deaths, none of them could prevent infection and block transmission for symptomatic case. The serum neutralizing antibody, either induced by natural infection or vaccination, seems to decrease quickly^[4]^. It is extremely eager and urgent to develop medications that can prevent COVID-19 infection.

In September 2022, COVID-19 broke out in the Inner Mongolia Autonomous Region, particularly in Hohhot. Due to an increase in COVID-19 patients, several designated COVID-19 hospitals and Fangcang shelter hospitals have been built in Hohhot. A number of medical teams from all over China joined these hospitals. However, these medical teams were at high risk for COVID-19 infection, and many members were infected within weeks.

The SA58 Nasal Spray is an intranasal spray developed by Sinovac Life Sciences Co., Ltd. The active ingredient of the SA58 Nasal Spray is a broad-spectrum neutralizing antibody with a high neutralizing capacity against different Omicron VOC sub-variants, such as BA.1, BA.2, BA.5, that can bind to the novel coronavirus, thus preventing the virus from entering host cells and blocking viral infection^[5-9]^.

The purpose of this study was to evaluate the effectiveness and safety of SA58 Nasal Spray in medical personnel who were working in designated COVID-19 hospitals and Fangcang shelter hospitals.

## Materials and Methods

### Study design and procedures

From October 30 to November 31, 2022, all medical personnel transiently working in 2 designated hospitals and 4 Fangcang shelter hospitals (alternate care sites) of COVID-19 in Hohhot, the Inner Mongolia Autonomous Region were involved in this study. This study is an open-label, blank-controlled study with the principles of compassionate medicines. All medical personnel who agreed to use SA58 Nasal Spray (drug group) were asked for informed consent, while the medical personnel who are NOT using the drug were divided into the blank control group. SA58 Nasal Spray should be used twice a day with an interval of 6 hours for about 30 days. The observation period was from Oct 31 to Nov 30, 2022, lasting for 30 days.

This study received approval from the Institutional Review Board of the Inner Mongolia Fourth Hospital (202223). This study is registered with ClinicalTrials.gov, NCT05664919.

### Selection criteria

All medical personnel with RT-PCR negative results for SARS-CoV-2 were enrolled, and the key exclusion criteria were history of allergic reactions, pregnant or unfit for intranasal administration.

### Investigational product

The SA58 Nasal Spray manufactured by Sinovac is a liquid external medicine containing 5 mg/ml of antibodies which has been verified to be able to neutralize many variants of SARS-CoV-2 PANGO lineage *in vitro*. The SA58 Nasal Spray is self-administered intranasally and is recommended to be given twice daily at 6-hour intervals.

### Effectiveness Assessment

As the medical personnel were not working at a fixed schedule in those hospitals and many of them were working on different shifts and rotations during the 30-day observation, the working person-day was mainly calculated and used for further analysis. Throat swab sampling and RT-PCR for SARS-CoV-2 are conducted for all participants every day. Positive cases who never used SA58 Nasal Spray or who did not use SA58 Nasal Spray 72 hours before sampling were considered as NOT using medication. Whereas positive cases that used SA58 Nasal Spray 72 hours before sampling were classified as using medication.

### Safety assessment

All the medical personnel who used SA58 Nasal Spray were asked to report adverse event via an APP of medication information collection in WeChat on a daily basis.

### Statistical analysis

The effectiveness of the SA58 Nasal Spray was mainly evaluated by calculating the cumulative incidence of COVID-19 cases. The cumulative incidence for each group was defined as the numbers of COVID-19 cases divided by the number of person-day during the observation.

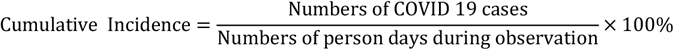

The effectiveness was evaluated with the following formula.

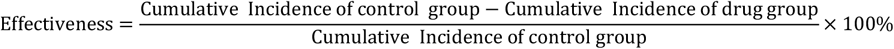

Statistical analysis was conducted using the SPSS 22.0 statistical package and SAS 9.4.

## Results

A total of 6,662 RT-PCR negative medical personnel working in the 2 designated hospitals and 4 Fangcang shelter hospitals receiving COIVD-19 cases in Hohhot, the Inner Mongolia Autonomous Region were enrolled in this study. Among the 3,368 participants using SA58 Nasal Spray, 1,736 participants reported their medication information via APP. The median age of the APP-reported participants was 34 years (ranging from 24 to 58 years), with approximately 23% in their twenties, 53% in their thirties, 21% in their forties and 2% in their fifties.. The gender ratio (M/F) was 23%/77%. No special coexisting condition was recorded among the participants.

Since the medical personnel were not working at a fixed schedule in those hospitals and many of them were working on different shifts and rotations during the 30-day observation, we counted the total working person-day as 135,544, amongst the person-day of in drug group was 27,103 and that of control group was 110,441 (Fig.1).

**Figure 1.**
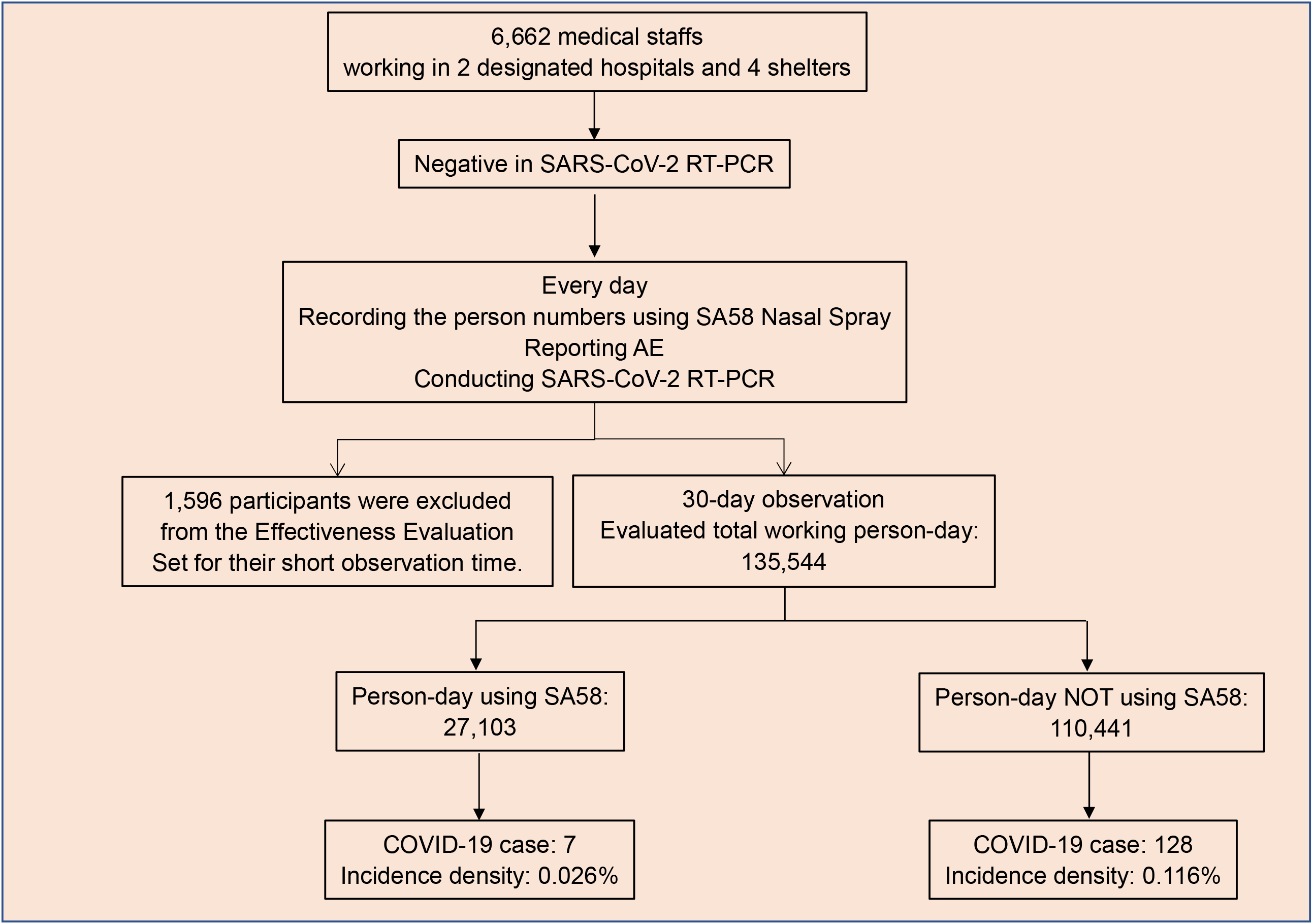
Study design and procedures of SA58 Nasal Spray in 6,662 medical personnel.

### Safety

Of the 1,736 medical personnel who reported the SA58 Nasal Spray medication information via APP, 1,794 AEs were reported from 497 medical personnel.. The incidence of AEs was 28.6% (497/1,736). The majority of AEs were administrative site AEs, including rhinorrhea (14.5%), nasal mucosal dryness (9.6%), sneezing (8.7%), nasal obstruction (6.0%), headache and dizziness (2.0%), pharyngolaryngeal discomfort (1.0%), discomfort in nose (0.9%), cough (0.5%), nausea (0.4%), expectoration (0.4%), rash (0.3%) were infrequently noticed (Fig. 2). Fever and other systematic AEs were extremely rare. The severity of all the AEs are mild, and all of them disappeared quickly without affecting daily working.

**Figure 2.**
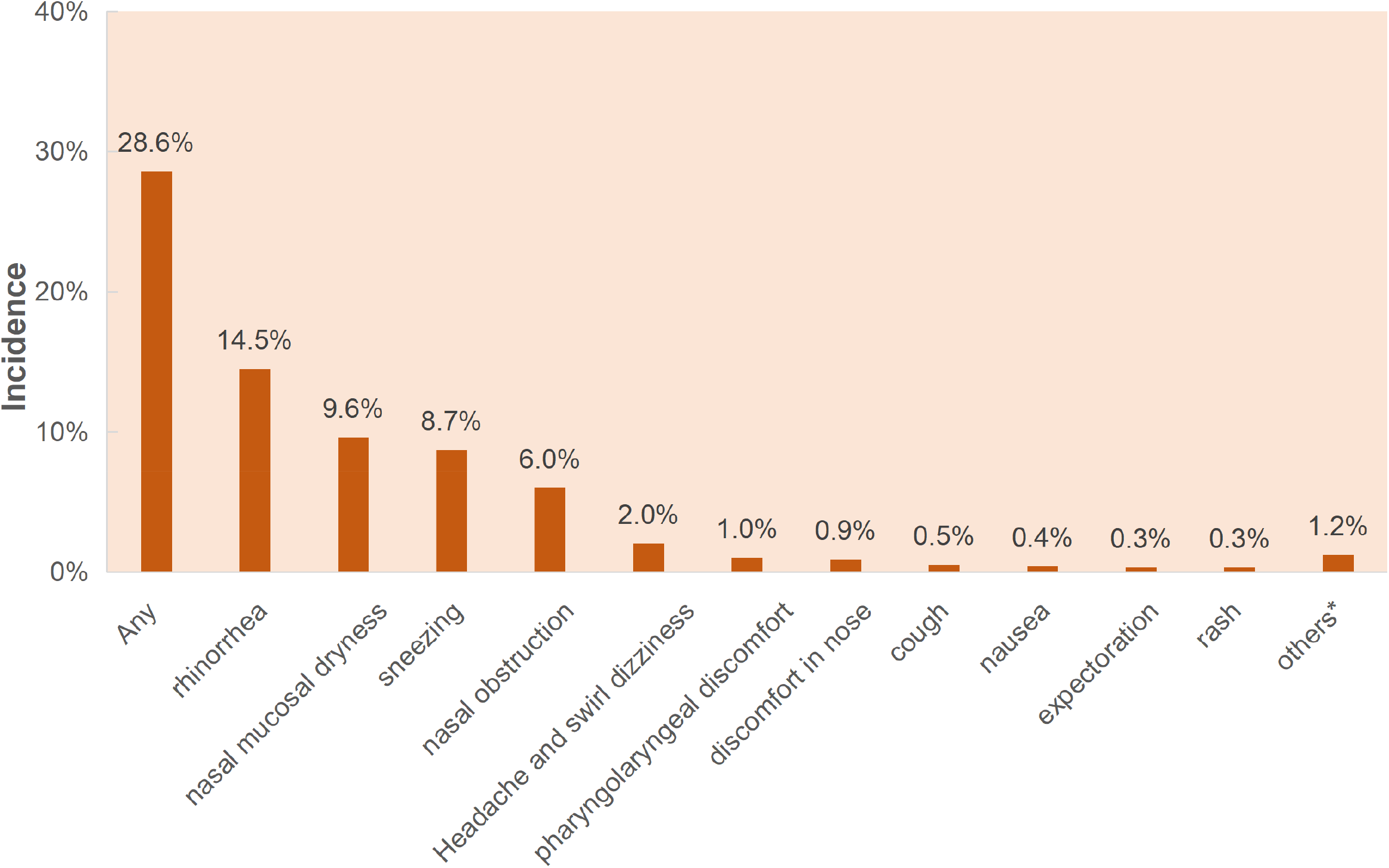
The reporting adverse events (AEs) incidence in the drug group during the 30-day observation. Various AEs are indicated in X-axis and the AEs incidence is indicated in Y-axis. *Others included mild fever, palpitation, muscle pain, bleeding nose, cold sweat, pruritus cutaneous, reaction gastrointestinal, eye pain, gasping, tight chest, dry mouth, redness of eyelid conjunctiva, skeletal pain

### Effectiveness

A total of 6,662 medical personnel were involved in this study: 3368 used SA58 Nasal Spray from the drug group, and 3294 not used from blank control group. During the observation period, 1,596 participants from the drug group no longer worked in the six hospitals within 2 days of receiving the SA58 Nasal Spray, so they were excluded from Effectiveness Assessment Set. There were 135 RT-PCR positive cases were found among 5,066 staff (1,596 participants excluded from the 6,662 medical personnel), with a crude infection rate of 2.66%. Of the 135 positive cases, 128 were in the control group, 7 cases in the drug group. The cumulative number of cases during the observation period showed a rapid increase of positive cases in the control group, whereas a remarkably slow increase in the drug group (Fig. 3). The cumulative incidence was 0.026% (7 cases out of 27,103 person-days) in the drug group and 0.116% (128 cases out of 110,441 person-days) in the control group. The effectiveness of the SA58 Nasal Spray for protecting infection in the medical personnel working in the COVID-19 designated hospitals and Fangcang shelter hospitals was calculated as 77.7% (95% CI: 52.2% - 89.6%).

**Figure 3.**
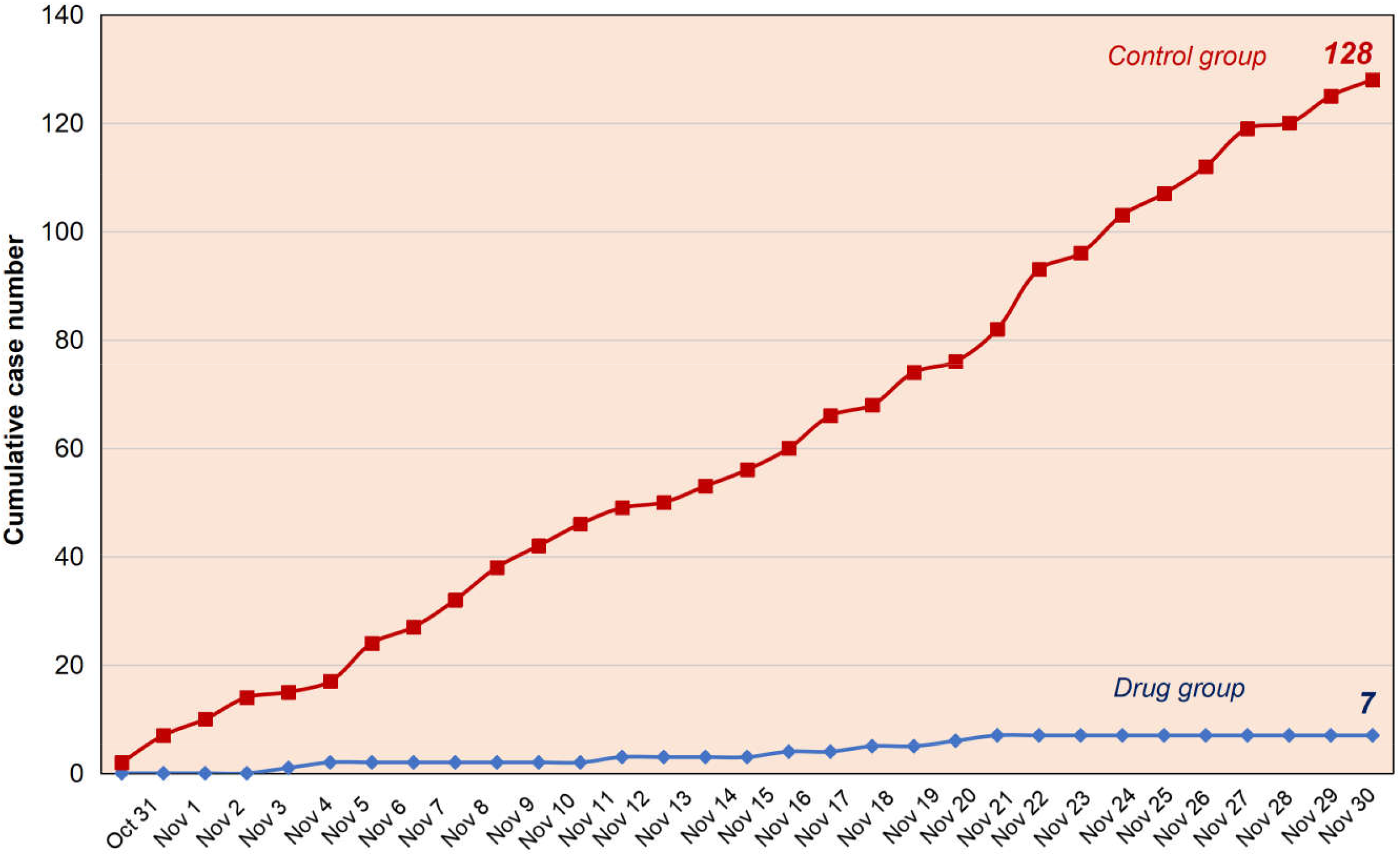
Cumulative COVID-19 cases in the drug group and the control group during 30-day observation. The dates are indicated in X-axis. Blue curve: drug group. Dark red curve: control group.

## Discussion

An open-label, blank-controlled study of SA58 Nasal Spray has been conducted among medical personnel working in designated hospitals and Fangcang shelter hospitals for COVID-19 during the period of October 31 to November 30, 2022. Virus genome sequencing assay showed that in Hohhot the Omicron VOCs circulating during this period were BF.7, BA.5.2, BA.5.2.1, BQ.1.2, etc., among which BF.7 was the predominant one. SA58 Nasal Spray showed farmable effectiveness. The 30-day observation period has witnessed a significantly lower incidence of COVID-19 cases in the drug group compared to the control group. The effectiveness of SA58 Nasal Spray for preventing SARS-CoV-2 infection among medical personnel working in the COVID-19 designated hospitals and Fangcang shelter hospitals as high as 77.7% (95% CI: 52.2% - 89.6%). The SA58 Nasal Spray was also shown to be well-tolerant. 497 out of 1736 participants reported AEs with an incidence rate of 28.6%. Considering the participants without AEs tended not to report their medical information via APP, the actual incidence of AEs could be lower than 28.6%. Among those participants who reported local AEs, most of them using SA58 Nasal Spray recorded that those mild AEs were almost ignorable and did not affect their daily work.

Infection and transmission of COVID-19 are not only attributed to virus variants, but also to many other physical components. It is worth emphasizing that all participants in this study are medical personnel dealing with COVID-19 cases. As a population of high risk, medical personnel usually suffer from a higher infection rate. Despite being equipped with eligible PPEs, the medical personnel in the control group reported a crude COVID-19 infection rate of 2.66% during the 30-day observation, which was remarkably higher than the infection rate in general population in Hohhot during that period. In addition to PPE, the SA58 Nasal Spray provides another specific protection tool that is easy to use for medical personnel and other high-risk professionals when in service.

Although Omicron VOC has shown a mild clinical severity in general, it still can cause severe clinical outcomes in the elderly, as well as the population co-existing with severe underlying diseases^[10]^. It is believed that it is extremely important to fully vaccinate these populations to avoid severe consequence^[11, 12]^, but that it is ineffective at preventing infection. Although currently there is still no SA58 Nasal Spray data on the elderly and those with underlying diseases, light local AEs from this study allow us to be confident about the safety in those populations.

We must recognize that the study here is not a placebo-controlled, observer-blinded study that may introduce data bias. Frequent rotations of medical personnel between different medical institutions make it difficult for many participants to complete all 30 days of observation. Additionally, it is also difficult to precisely monitor the daily frequency of medications. The protective effectiveness of SA58 Nasal Spray remains to be verified by more clinical trials.

In summary, this clinical study of the SA58 Nasal Spray on medical personnel showed good tolerance and good effectiveness for preventing COVID-19 infection, suggesting further application in other population in the real world.

## Data Availability

All data produced in the present study are available upon reasonable request to the authors.

## Author Contributions

Y. Cao, S. Zhang, and Y. Hu: study design; X. Dong: manuscript drafting; S. Si, and C. Jin: stud performance; S. Si, J. Li, and L. Fang: data analysis; F. Xue, and G. Zeng: provided editorial assistance before submission; X. Xie, B. Kan, and Y. Cao: revised it critically for important intellectual content. All authors have read and agreed to the published version of the manuscript.

